# Mapping the Shadows: A Comprehensive Exploration of Breast Ironing’s Effects on Women and Girls through a Systematic and Scoping Review

**DOI:** 10.1101/2024.06.24.24309436

**Authors:** Benjamin Olusola Ajibade, Prince Ubah, John Kainesie, Umma Suleiman, Maxwell Omoruyi, Adediran-Ibegbunam Tolulope Olubunmi

## Abstract

**Background:** The research paper titled "Mapping the Shadows: A Comprehensive Exploration of Breast Ironing’s Effects on Women and Girls through a Systematic/Scoping Review" examines the cultural practice of breast ironing (BI) and its significant impact on the physical, psychological, and social well-being of women and girls. It explores the biopsychosocial implications of breast ironing on women, assesses knowledge and attitudes towards the practice, and analyses its prevalence and consequences.

**Methods:** This study employs a systematic/scoping review design following the PRISMA guidelines to collect and analyse qualitative and quantitative data. Data collection involved a comprehensive search of online databases, including EBSCO, PubMed, Google Scholar, and others. Four investigators conducted independent searches using specific keywords related to breast ironing. The inclusion criteria focused on studies examining the biopsychosocial impact of breast ironing within the past 10 years. The data were analysed using thematic analysis, chi-square tests, and logistic regression models.

**Results:** The practice of breast ironing results in severe physical and psychological trauma, including pain, tissue damage, infections, abscesses, and breast cancer. Psychologically, victims experience anxiety, depression, low self-esteem, and PTSD. The study found significant barriers to healthcare access due to stigma and lack of awareness. The prevalence of breast ironing is highest in the Littoral region (53%), with varying rates across other regions. Chi-square tests revealed significant relationships between BI and health outcomes, such as severe pain and family health issues (p-value < 0.001). Logistic regression indicated a strong association between BI exposure and negative health outcomes.

**Conclusion:** Breast ironing is a harmful cultural practice with substantial negative impacts on the health and well-being of women and girls. Comprehensive interventions, including legal measures, community education, and support services, are crucial to eradicate this practice and protect the rights and health of affected individuals.

## Background

Breast Ironing (BI) persists as a deeply rooted cultural practice with potentially detrimental effects on the physical, psychological, and emotional well-being of its victims. The practice of BI involves physically compressing the breasts of young girls through several crude means, primarily aimed at hindering breast development, impeding early sexual interest and debut, hindering adolescent pregnancies, enabling extended education, and safeguarding chastity for marriage. ^1, 2^

Although the historical genesis of BI is difficult to establish, it is typically transferred from mothers to their daughters ^3^. In regions where BI is practised, it is regarded as a direction for appropriate behaviour and treatment of girls ^4^. Despite the intention to delay sexual initiation and protect pubescent girls, the harmful short-term and long-term magnitudes of BI on the victims’ well-being significantly outweigh its purported benefits ^4^. BI is connected to various health risks and negative outcomes, such as adverse physical health effects ^3, 5, 6, 7^.

Subsequently, several advocacy groups, specifically NGOs and the United Nations classify BI as a harmful practice and a violation of the rights of the girl-child, advocating for its eradication. Nonetheless, the efforts to prevent this act have encountered resistance in communities that adhered to this practice ^8^. The insight of BI remains limited. A study by Ngunshi ^3^ aimed to investigate BI’s intricacies, motivations, historical context, and cultural underpinnings, suggesting that it might have originated as an ancient breast massage practice to address uneven breast growth. Its primary intent is to safeguard young girls from early sexual experiences and associated risks. BI uses various heated objects for this practice, and typically, the perpetrators are familial figures ^3^.

Breast ironing affects an estimated 3.8 million women worldwide, with the majority of cases occurring in Africa, and it has been identified as one of the five under-reported crimes related to gender-based violence ^9^. This practice is most commonly performed by a family member, particularly mothers, in 58% of cases. In Cameroon, it is estimated that between 25% and 50% of girls undergo breast ironing. Additionally, around 1,000 girls aged 9 to 15 in the UK are currently thought to be at risk of this practice ^10^.

In a 2016 debate in the House of Commons, Berry ^11^ discussed breast ironing (BI) while addressing the awareness and legislative journey against Female Genital Mutilation (FGM) in the UK. The debate highlighted the lack of prosecutions despite significant legal efforts and drew parallels with the lesser-known practice of breast ironing ^12^. Both practices are rooted in cultural beliefs aimed at controlling female sexuality, necessitating increased awareness, legal frameworks, and support systems to protect affected girls. Lazareva ^13^ found, through interviews with Margaret Nyuydzewira, head of the diaspora group CAME Women and Girls Development Organisation ^14^, that at least 1,000 women and girls in the UK had been subjected to BI intervention. It was concluded that no systematic study or formal data collection exercise on the topic had been conducted in the UK. It was also noted that BI is typically performed in the UK rather than abroad, unlike FGM, with about 15 to 20 recent cases reported in Croydon alone.

Although several studies have explored BI’s impact on women and pubescent girls, most have been concentrated in Cameroon, despite indications that BI is prevalent in other African countries and is becoming a global issue. These studies have mostly focused on short-term experiences or advocated against BI ^2, 3, 15, 16^. Some research has investigated the practice’s methods, motivations, outcomes ^7^, long-term health-related consequences, and victims’ quality of life ^6^.

### Motivations Behind BI Practice

Contrary to other "harmful traditional practices," such as female genital mutilation or child marriage, BI is driven by the intention to safeguard young girls from the perils of adulthood, early sexual experiences, pregnancies, and early marriages. It also strives to promote future education and well-being. BI is rooted in embedded sociocultural attitudes, traditional norms, and gender inequality. Many patriarchal societies perpetuate these norms, aiming to maintain gender roles and privileges, favouring men over women. BI seeks to extend the period during which girls can prepare for adulthood and marriage, thereby postponing pregnancy, facilitating education, and delaying marriage to enable the realisation of individual potential ^17, 18^. Ethnographic research by Pemunta ^17^ reveals BI as a societal and disciplinary phenomenon to preserve societal norms and ensure the well-being of girls. Eriksson^18^ finds BI to hold diverse meanings in lived experiences.

### Physical Health Consequences of BI

BI inflicts negative physical impacts on victims. Breast pain, distinct from cyclical pain, is a common outcome ^1^. Other physical detriments comprise a violation of physical integrity, risk of severe pain ^19^ or discomfort, swelling, tissue damage, burns, scars, bruises, deformities, and discolouration on breasts ^2, 6, 7, 20, 21^. Research even links BI to traumatic breast injuries. Foley et al. ^22^ highlight long-term consequences, including scarring and psychological distress from burns. Frostbite-induced breast injuries are also documented to occur while using faulty cryotherapy or persistent cold exposure to soft tissue ^23^.

### Psycho-Social Health Consequences of BI

Nkwelle’s ^6^ study on long-term outcomes of BI in Cameroon underscores victims’ perceptions of physical, psycho-social, and emotional outcomes, such as severe pain, breast scars, stress, feelings of inferiority, sadness, and negative changes in quality of life. BI can contribute to emotional and psychological challenges, such as fear, low self-esteem, depression, shame, and mistrust in caregivers^24, 25^.

### Impact of BI Trauma on Health Outcomes

Though minimal research addresses BI’s long-term impact, related pain studies indicate potential physical and psychological trauma. Survivors of traumatic incidents, like the station nightclub fire, experience chronic pain, depression, and compromised quality of life ^26, 27, 28^. BI’s combined effects could potentially erode physical function and quality of life ^27, 29, 30, 31^.

### Effects of BI on Quality of Life

BI constitutes a traumatic event with repercussions on survivors’ quality of life (QOL). Trauma stemming from violence, accidents, or abuse correlates with lifestyle changes, decreased QOL, and coping difficulties, especially in women ^32, 33, 34, 35, 36^. This systematic review aims to aggregate and evaluate the findings of studies on BI, facilitating informed decision-making.

### Research Aim

This research aims to comprehensively explore the biopsychosocial impact of breast ironing on women.

### Research Objectives

1. To assess the knowledge levels of women regarding breast ironing.
2. To examine the attitudes of women towards the practice of breast ironing.
3. To investigate the prevalence and practices of breast ironing among women.
4. To analyse the physical, psychological, and social consequences of breast ironing on women.

### Research Question

What are the multifaceted biopsychosocial effects of breast ironing on women?

## Method

We conducted a systematic and scoping review using the PRISMA (Preferred Reporting Items for Systematic Reviews and Meta-Analyses) 2020 guidelines ^37^. We aimed to explore the knowledge, attitudes, and practices of breast ironing among research participants. To achieve this, we designed a tailored protocol; however, we are unable to register it on PROSPERO because the search had already been conducted. Our comprehensive search encompassed both published and grey literature, utilising a range of online databases, including American Premier (EBSCO), PubMed/Medline, Google Scholar, Embase, Web of Science, PsycINFO, and CINAHL. Given the scarcity of research on this topic, we considered all relevant publications to date.

### Search Strategy

Four investigators independently conducted the search for articles from August 1, 2023, to November 25, 2023. The search involved specific keywords such as "breast ironing," "breast flattening," "breast pounding," "chest ironing," "breast trauma," "breast injury," "knowledge," "attitude," "perceptions," "experiences," "views," "effects," "impact," "quality of life indicators," "health-related consequences and outcomes," "questionnaire," "interview," and "survey." These terms were manipulated using Boolean operators (AND, OR, NOT) to ensure a comprehensive search ^38^.

See S2 Table for Boolean search teams.

Additionally, we conducted a manual search by examining the reference lists of relevant papers. The search extended to other databases, including the World Health Organization (WHO), British Medical Journals (BMJ), and relevant policy documents and guidance such as NICE and library catalogues. We also included articles identified from the bibliographies of studies obtained from hand-searched journals.

Our review was limited to articles written in English and included various study designs that reported quantitative, qualitative, literature scoping and report of breast ironing. We applied strict inclusion and exclusion criteria to identify relevant articles. Our approach involved developing appropriate search terminologies for the topic, executing a search strategy using various search engines, critically evaluating selected articles, and extracting essential information from them without restriction to the year. However, a restriction was applied to the search years by the authors from 2014 to 2023, yielding a total of 1,740 publications. After removing duplicates, 1,386 publications remained. Further screening by titles and abstracts (857) reduced this number to 529 articles. Irrelevant articles (peer-reviewed and full text) were discarded (508), narrowing the selection to 21 articles.

See Fig 1 and S1 Table

**Fig 1.**
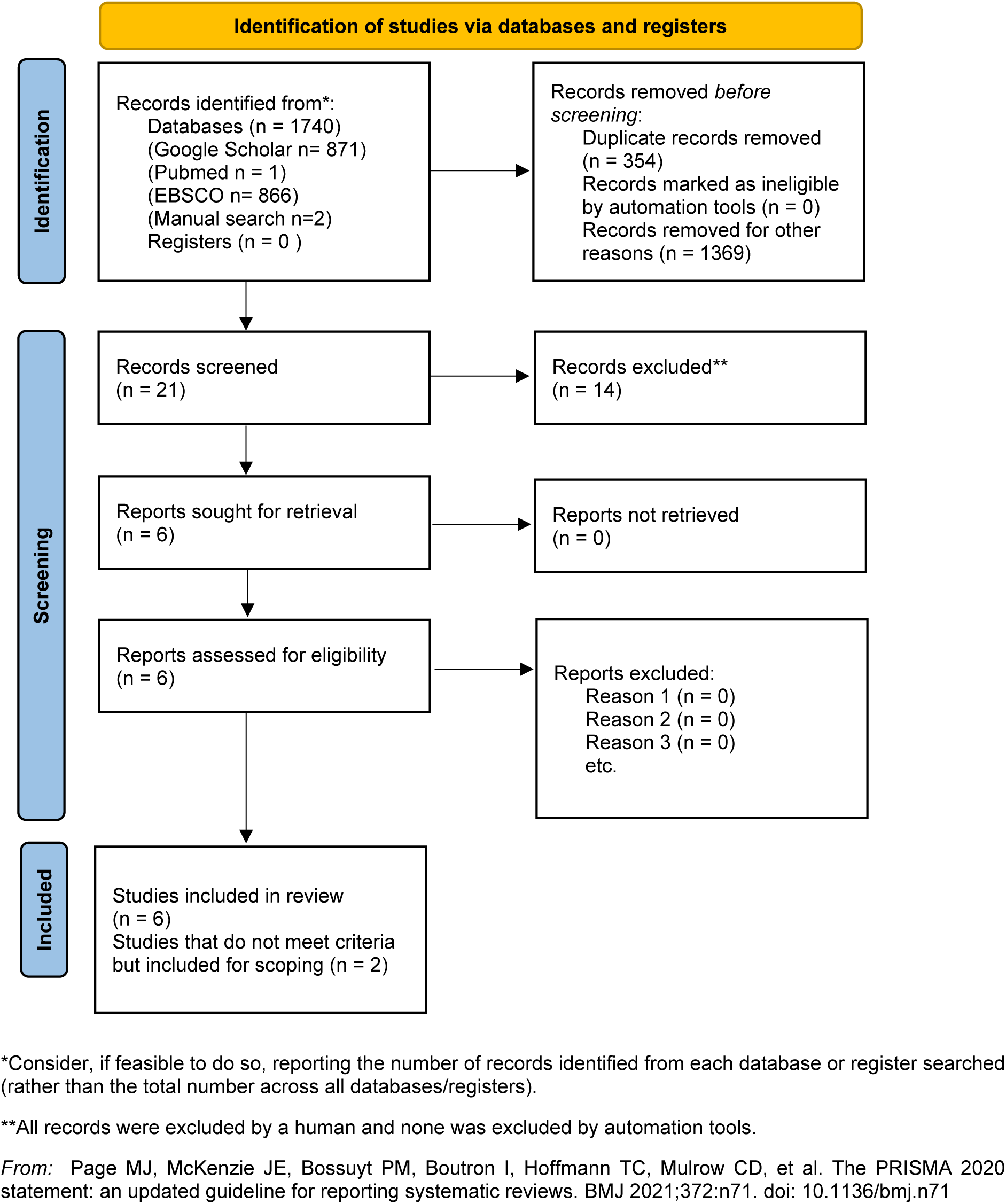
PRISMA 2020 flow diagram for new systematic reviews which included searches of databases and registers only.

The 21 remaining articles were further reviewed and screened by title, abstract, full text, and reference lists. Seven articles were excluded. Full-text articles were assessed for eligibility, with three excluded for not meeting quality criteria. Ultimately, 14 publications were included for a full assessment, but only 2 articles ^17, 39^ passed the final quality review, while 4 met some of the assessment criteria ^6, 7, 40, 41^ but were selected due to very limited work carried out in this area.

### Inclusion and Exclusion Criteria

#### Inclusion Criteria

Articles reporting on studies conducted within the past 10 years that examine the biopsychosocial impact of breast ironing on women, using quantitative or qualitative research methods. Case studies were included due to the limited number of primary research articles in this area.

#### Exclusion Criteria

Articles not written in English, studies conducted outside the specified timeframe, and research focusing solely on men or other populations.

By meticulously applying these inclusion and exclusion criteria, we identified relevant articles that provide a comprehensive understanding of the biopsychosocial effects of breast ironing on women using the PEO criteria ^54^.

PEO table.

| PEO | INCLUSION CRITERIA | EXCLUSION CRITERIA |
| --- | --- | --- |
| POPULATION | Women with breast ironing, | Women who did not undergo breast ironing, women with |
| EXPOSURE | Breast ironing, | Female genital mutilation |
| OUTCOME | Biopsychosocial impact of breast ironing | Others unrelated outcome |
| TYPES OF STUDY | Primary studies like qualitative research, quantitative study |  |

### Quality Assessment

Quality assessment in this study involved evaluating the methodological rigor and validity of the primary studies included. The process encompassed a thorough examination of each study’s design, sampling methods, data collection techniques, and analytical procedures. This ensured that the findings were reliable and that any potential biases were minimised. Each study was rated based on predefined criteria, which included aspects like the clarity of research questions, the appropriateness of the methodology, the robustness of the data analysis, and the overall consistency of the results. We conducted a systematic review involving 6 primary research studies, 2 articles ^17, 39^ passed the final quality review, while 4 met some of the assessment criteria ^6, 7, 40, 41^ but were selected due to very limited work carried out in this area. The other 2 papers included in the scoping review are one report and a literature review ^3, 43^. The quality of the studies included in this systematic/scoping review was assessed using criteria adapted from established quality assessment frameworks, specifically the Critical Appraisal Skills Programme (CASP) for qualitative studies to evaluate the credibility, relevance, and robustness of qualitative research ^44^ CASP Checklist Mixed Methods Appraisal Tool (MMAT), version 2018 ^45^ and Tulder’s ^46^ assessment checklist for quantitative studies. The assessment criteria focused on several key aspects to ensure a thorough evaluation of each study’s methodological rigor. See S4 to 6 Tables.

### Data Extraction

Data extraction involved systematically collecting relevant information from the selected studies. This process was guided by a standardised form to ensure consistency and comprehensiveness ^47^. Key data points included study objectives, population characteristics, interventions, outcomes, and significant findings. Extracted data were then organised into a database, facilitating easy comparison and synthesis across studies. This step was crucial for identifying patterns and drawing meaningful conclusions about the impact of breast ironing. See S3 Table

### Study Characteristics

The studies included in this review varied in terms of their geographical locations, sample sizes, and research methodologies. Most studies were conducted in regions where breast ironing is prevalent, such as Cameroon and parts of West Africa ^3, 6, 40^. The sample sizes ranged from small, qualitative studies involving in-depth interviews to larger, quantitative surveys. Research methodologies included ethnographic studies, case studies, cross-sectional surveys, and longitudinal analyses. These diverse study designs provided a comprehensive understanding of the practice and its impacts.

### Risk of Bias

Assessing the risk of bias was a critical component of this review. Each study was evaluated for potential biases that could affect the validity of the findings. Common sources of bias included selection bias, recall bias, and researcher bias. Strategies to mitigate these biases were noted, such as the use of random sampling, validated measurement tools, and triangulation of data sources ^48^. Studies with a high risk of bias were scrutinised carefully, and their findings were interpreted with caution to avoid drawing erroneous conclusions.

### Validation of Outcome Measures

Validation of outcome measures involved ensuring that the tools and methods used to assess the impacts of breast ironing were reliable and valid. This included verifying that the instruments used for data collection were appropriate for the cultural context and that they had been previously validated in similar settings. The consistency and reproducibility of the results were also considered, ensuring that the findings were robust and could be replicated in future studies ^44, 45, 46^.

### Ethics

Ethical considerations were paramount in this research. The studies included adhered to ethical standards such as obtaining informed consent from participants, ensuring confidentiality, and minimising harm. The sensitive nature of the topic required researchers to handle data with care and respect the dignity and autonomy of the participants. Ethical approval from relevant institutional review boards was a prerequisite for inclusion in this review. The researchers also addressed ethical dilemmas related to studying harmful traditional practices, ensuring that the research contributed to broader efforts to protect and promote human rights ^48^.

By meticulously addressing these aspects, the study provides a comprehensive and credible assessment of the impact of breast ironing, contributing valuable insights to the field and informing interventions aimed at eradicating this harmful practice.

## RESULTS

### Falana TC. (2020) ^40^ “Breast Ironing: A Rape of the Girl-Child’s Personality Integrity and Sexual Autonomy"

Falana’s ^40^ article investigates the harmful practice of breast ironing, or breast flattening, and its impact on the personal integrity and sexual autonomy of young girls. Through qualitative research and expository analysis, the study reveals the clinical, psychological, and social consequences of this practice, which involves using heated objects to retard breast development. Prevalent in parts of Cameroon, breast ironing is performed under the pretext of protecting girls from sexual harassment, rape, early pregnancy, and dropping out of school. Culturally, it is seen as a way to preserve chastity and prevent premarital sex, compelling mothers to carry out the practice on their daughters. The physical effects include breast tissue damage, infections, and long-term deformities, while the psychological effects encompass trauma, reduced self-esteem, and long-lasting mental health issues. Socially, breast ironing reinforces gender-based violence and perpetuates the cycle of abuse and subjugation of women.

Falana^40^ frames breast ironing as a severe violation of human rights, comparing it to rape due to its lack of consent and violent nature. The article calls for the recognition of breast ironing as a form of gender-based violence that infringes on girls’ rights to security, dignity, and freedom. The author advocates for strict laws and penalties to abolish the practice and urges global condemnation and concerted efforts to end it. Emphasising the need for awareness and education on the rights of the girl-child, Falana’s ^40^ article provides a detailed examination of breast ironing, highlighting its severe consequences and cultural underpinnings. The article argues for urgent action to address and eradicate this harmful practice to protect the rights and well-being of young girls, aiming to foster a societal shift towards recognising and upholding their sexual autonomy and personal integrity.

### Gorar M (2022) ^43^ "Breast Ironing in the UK and Domestic Law"

The literature examines the practice of breast ironing, a form of gender-based violence, and its legal implications in the UK. Breast ironing involves massaging or pressing the breasts of young girls to delay development, aiming to protect them from sexual harassment and early marriage. Predominantly practised in certain African communities, it has been reported among diaspora communities in the UK. The practice leads to severe physical and psychological consequences, including tissue damage, pain, and trauma, infringing on the victims’ physical and sexual autonomy.

While there is specific legislation against FGM in the UK, breast ironing lacks explicit legal recognition, presenting gaps in the law and challenges in prosecuting related cases. Classified under the broader spectrum of gender-based violence, breast ironing is considered a form of domestic violence and honour-based violence. The author advocates for the explicit legal recognition of breast ironing as a criminal offence, increased awareness and education within communities, and enhanced support mechanisms for victims.

The true prevalence of breast ironing in the UK is likely underreported due to cultural sensitivities and lack of awareness, posing challenges for law enforcement and social services. Significant legal loopholes allow the practice to persist unchallenged, highlighting the need for a stronger legal framework. The persistence of breast ironing within certain communities underscores the necessity for culturally sensitive interventions, collaboration with community leaders, and culturally competent education programs to change attitudes and practices.

Mukaddes Gorar’s ^43^ article highlights the critical issue of breast ironing and its overlooked status in the UK’s legal framework. The practice’s severe impact on girls’ physical and psychological well-being, coupled with legal gaps, calls for urgent attention and action. Enhanced legal measures, awareness campaigns, and supportive services for victims are essential steps toward eradicating this form of gender-based violence.

### Ngunshi BR. (2011) ^3^ "Breast Ironing: A Harmful Traditional Practice in Cameroon"

The report examines the practice of breast ironing, which involves flattening young girls’ developing breasts using heated objects to delay puberty and protect them from sexual harassment and early pregnancy. This practice affects women across all 10 provinces of Cameroon, with the highest prevalence in the Littoral province (53%), particularly in urban areas like Douala, and the lowest in the northern, predominantly Muslim regions (7%).

Breast ironing is primarily carried out by mothers (58%), followed by nannies (10%), sisters (9%), and grandmothers (7%). Girls showing signs of puberty before the age of nine are twice as likely to undergo breast ironing. The rationale behind this practice includes protecting girls from sexual harassment and rape, preventing early pregnancy and marriage, and allowing them to pursue education.

The heated objects used include spatulas (24%), stones (20%), pestles (17%), and other items like coconut shells and wooden spoons. Breast ironing can cause severe pain, tissue damage, infections, deformities, and psychological issues, including abscesses, cysts, severe chest pain, infections, malformed breasts, and even breast cancer. Despite its harmful effects, the practice is seen by perpetrators as a protective measure, with mothers believing they are safeguarding their daughters’ futures.

Statistical data shows higher prevalence rates in urban areas compared to rural ones. The likelihood of undergoing breast ironing is 50% for girls developing breasts before age nine, decreasing to 38% before age 11, 24% before age 12, and 14% before age 14. Objects used include hot wooden spoons or brooms (24%), stones (20%), pestles (17%), breast bands (10%), and other items like hot fufu or hot seeds (15%).

Gender Empowerment and Development (GeED) recommends increasing public awareness of the harmful effects of breast ironing and comprehensive sex education to address misconceptions about puberty and pregnancy. Strengthening mechanisms for redress and support for victims, documenting women’s rights violations, and advocating for legislative changes are also essential. The practice of breast ironing is a severe form of gender-based violence prevalent in Cameroon due to deep-rooted cultural norms and misconceptions. Efforts to combat this practice must include raising awareness, educating communities, supporting victims, and implementing stringent legal measures. The research underscores the need for a multi-faceted approach to eradicate breast ironing and protect the rights and well-being of young girls in Cameroon.

### Nkwelle NNN. (2019) ^6^ "The Long-Term Health-Related Outcomes of Breast Ironing in Cameroon"

The study surveyed 230 women who experienced breast ironing (BI) in Cameroon. Participants’ ages ranged from 18 to 80 years, with the majority in the 18-40 age group. The practice was more prevalent in the Southwest, Northwest, Littoral, West, and Center regions, with higher prevalence among ethnic groups such as Duala, Manyu (Banyan, Ejagham, and Akwaya), Eton, Bamelike, and Bafut. Most participants reported multiple instances of BI during their adolescence.

Key findings include significant long-term physical, psychological, and emotional health-related outcomes associated with BI. Physically, severe pain, breast scars, abscesses, and burns were common. Psychologically, high levels of stress, feelings of inferiority, and depression were prevalent. Emotionally, many women reported sadness, frustration, and shame. Negative changes in quality of life (QOL) were strongly associated with the BI experience, impacting physical health through persistent pain and discomfort, psychological health through long-term trauma affecting self-esteem and social interactions, and social health by straining marital and family relationships.

Chi-square tests revealed significant relationships between BI and various health outcomes: severe pain and marital/family health (p-value < 0.001), breast scars and frequent pain (p-value < 0.001), stress and feeling inferior (p-value < 0.001), and sadness and pain (p-value < 0.001). Logistic regression models indicated the odds of experiencing negative health outcomes based on BI exposure, with lower odds (OR < 1) for experiencing breast scars, frustration, and shame compared to those without these experiences, and significant predictors of long-term emotional and psychological health issues like depression and self-esteem.

The study highlights the severe and long-lasting impact of BI on women’s health in Cameroon, emphasizing the need for public health interventions and advocacy to eradicate the practice and support the affected women.

### Nyangweso M. (2022) ^41^ "Contending with Health Outcomes of Sanctioned Rituals: The Case of Puberty Rites"

The research provides a detailed analysis of the health outcomes associated with puberty rites. The study surveyed 60 respondents, using both primary data from in-depth interviews and focus group discussions, and secondary data. The respondents included 5 doctors, 5 social workers, 30 survivors of FGM/C and breast ironing, and 20 other individuals knowledgeable about these practices, with participants from Kenya and the African diaspora.

Key findings highlight significant health outcomes related to puberty rites. Breast ironing is linked to severe pain, cysts, abscesses, infections, milk or fluid discharge, cancer, psychological problems, and sometimes the complete disappearance of breasts. FGM/C is associated with immediate effects like severe pain, urine retention, shock, hemorrhage, and infection, and long-term effects such as genital damage, cysts, abscesses, keloids, scarring, dyspareunia, childbirth complications, and sexual dysfunction.

The social and psychological impacts include high levels of anxiety, terror, humiliation, betrayal, and significant psychological stress, leading to conditions similar to PTSD. These practices are deeply embedded in cultural and religious norms, justified as means to control female sexuality, ensure chastity, and maintain family honor.

A survey of 113 individuals assessed awareness and opinions about FGM/C in industrialized countries. Results showed 60% awareness of its practice, 70% knowledge of its illegality among immigrants, 81% viewing it as a human rights violation, 25% believing religion recommends it, and 81% advocating for its eradication.

The health implications of these practices are significant, with short-term effects including haemorrhage, severe pain, urinary tract infections, injury to adjacent tissues, death, fractures or dislocation, and haematocolpos, and long-term effects such as keloid formation, vulval abscess, sexual dysfunction, pregnancy and childbirth complications, psychological damage, penetration problems, and lack of sexual response.

The study concludes that puberty rites have profound health and social impacts, requiring a holistic and intersectional approach that considers the cultural and religious contexts of the affected communities. Emphasising the need for medical training to handle the health outcomes of these practices is essential to improve care and support for survivors.

### Oniyangi SO. and Tosin JA. (2022) ^39^ "Influence of Cultural Practices on the Health of Yoruba People Living in Ilorin South LGA, Kwara State"

The research examines the health impacts of cultural practices such as breast ironing, puerperal baths, and nutritional taboos among the Yoruba people. The study used a descriptive research design with a survey method, involving 200 respondents selected through multi-stage sampling. A researcher-designed questionnaire with a reliability coefficient of 0.75 was used to collect data, which were analysed using frequency counts, percentages, and chi-square tests at a 0.05 alpha level.

The findings reveal that 88.6% of respondents acknowledged negative health impacts from breast ironing, which causes swelling, burning, irritation, increased risk of breast cancer, fever, extreme pain, and delayed breast growth. The chi-square analysis confirmed a significant negative influence with a calculated value of 532.24, exceeding the critical value of 16.92. Regarding puerperal baths, 73.4% of respondents recognized negative health impacts, such as increased body temperature, risk of perinatal cardiac failure, fever, and burns, with a chi-square value of 259.16, also above the critical value. Nutritional taboos were noted by 92.8% of respondents to have negative health impacts, leading to severe malnutrition, immune system ineffectiveness, and slowed physical growth due to nutrient shortages. This was confirmed with a chi-square value of 780.24.

The descriptive statistics indicate a high awareness of the negative impacts of these cultural practices, with significant agreement among respondents. For breast ironing, 51.8% strongly agreed and 36.8% agreed on its negative impacts. For puerperal baths, 43.6% strongly agreed and 29.8% agreed. Nutritional taboos had 63.7% strongly agreeing and 29.1% agreeing on their negative effects.

The authors recommend abolishing breast ironing and educating young girls on sexual values to avoid premarital sex, modifying puerperal baths to use slightly cool water for new mothers, and revising nutritional taboos to ensure essential nutrients are not restricted for vulnerable groups like invalids, pregnant and lactating mothers, and infants. They also suggest promoting healthy lifestyles to end harmful socio-cultural practices.

The study concludes that these cultural practices have significantly negative effects on the health of the Yoruba people in Ilorin South LGA, highlighting the need for cultural education and health promotion to mitigate these harmful effects.

### Pemunta NV. (2016) ^17^ "The Social Context of Breast Ironing in Cameroon"

The research explores the ethnographic and social factors underpinning the practice of breast ironing. The study estimates that 1.3 million Cameroonian girls are victims of breast ironing, with the practice also observed among the Cameroonian and African diaspora in countries such as Britain. The prevalence is highest in the Littoral region (53%), followed by the West and Centre (31%), Adamawa (30%), Northwest (18%), East (17%), South (14%), Southwest (11%), and lowest in the North and Extreme North regions (7%).

Breast ironing causes severe physical effects such as abscesses, cysts, infections, tissue damage, and sometimes the disappearance of one or both breasts. Other severe outcomes include breast cancer and psychological trauma. Victims often suffer from low self-esteem, emotional distress, anxiety, depression, and other psychiatric disorders. The practice is seen as a disciplinary technique to protect young girls from sexual harassment, rape, and unwanted pregnancies, aiming to make girls’ bodies unattractive to men and shield them from the sexualized male gaze. It is conceptualized as a means of governing the female body within a patriarchal society and is passed down through generations.

Factors such as the HIV/AIDS pandemic and high rates of under-aged pregnancy have led to the reinterpretation of breast ironing as a protective measure. Urbanization and technology have also contributed, with urban-based mothers more likely to practice breast ironing due to fears of increased exposure to sexual abuse. The anonymity of urban life and the influence of technology have weakened traditional mechanisms of social control.

The study used an ethnographic approach with in-depth interviews, focus group discussions, informal discussions, and participant observation. The sample included 16 girls subjected to breast ironing, 5 activists, 6 practitioners (mothers who performed breast ironing), and other informants selected through snowball and purposive sampling. Data were collected through tape-recorded interviews and notes, ensuring reliability and validity through a combination of data gathering methods. Significant negative health impacts were corroborated by qualitative data, revealing patterns and differences through content analysis.

The study recommends empowering girls and women through health enhancement interventions, pregnancy-prevention programs, and access to sexual and reproductive health services. It emphasizes the need for education and rights, ensuring unfettered access to education for young girls, even when pregnant, and investing in human capital. The creation of youth-friendly contraceptive services and reproductive health centers tailored to adolescents’ needs, and the integration of comprehensive sex education into secondary school curricula are also suggested.

In conclusion, breast ironing is a harmful cultural practice with significant physical and psychological health consequences. The study highlights the need for comprehensive interventions to protect young girls’ well-being and ensure their rights to education and health.

### Tapscott R. (2012) ^7^ "Understanding Breast Ironing: A Study of the Methods, Motivations, and Outcomes of Breast Flattening Practices in Cameroon"

The research explores the cultural practice of breast ironing. The practice affects about a quarter of all girls and women in Cameroon, with prevalence rates varying by region: Littoral (53%), West and Center (31%), Adamawa (30%), Northwest (18%), East (17%), South (14%), Southwest (11%), and North and Extreme North (7%).

Breast ironing is most often performed by the girl’s mother (58%), but also by nurses, caretakers, aunts, older sisters, grandmothers, and occasionally the girl herself or other family members. Common tools include grinding stones, wooden pestles, spatulas, brooms, belts, leaves, napkins, plantain peels, stones, fruit pits, coconut shells, salt, and ice. The practice varies in intensity and duration, ranging from a single ceremonial treatment to repeated sessions over weeks or months.

The primary motivation for breast ironing is to deter unwanted sexual attention by delaying breast development, protecting girls from early pregnancies, and ensuring they can continue their education without the distractions of early sexual activity or motherhood. It reflects broader gender norms and power dynamics, including the control of female sexuality and reproductive rights.

The health impacts are severe. Physically, it can cause swelling, burning, irritation, pimples, abscesses, fever, and extreme pain. Long-term effects include overgrowth or stunting of breast development, difficulty breastfeeding, scarring, and increased risk of breast cancer. Cases of second-degree burns requiring medical treatment, including skin grafts, have been reported. Psychologically, many girls experience distress, fear, shame, internalized blame, and social exclusion, which can lead to anxiety, depression, and a loss of self-confidence. Socially, the practice is often hidden, with many women not associating their physical or psychological issues with their experiences of breast ironing until much later in life.

The practice is deeply entrenched in cultural beliefs and traditional practices, making it difficult to eradicate. Limited access to sex education, contraception, and legal protections against sexual exploitation exacerbates reliance on breast ironing as a protective measure. As women gain access to education and career opportunities, there is increased pressure to delay marriage and motherhood, and breast ironing is viewed as a way to navigate these changing social conditions.

To address breast ironing, the study recommends raising public awareness about its harmful effects and educating communities about human biology. Advocacy efforts by NGOs should be supported, and there should be a push for the enforcement of legal instruments that protect women’s and children’s rights. Community engagement, including men, women, boys, and girls in both rural and urban areas, is essential to create sustainable change.

Breast ironing is a harmful cultural practice with significant physical and psychological health impacts. Addressing this issue requires a comprehensive approach that includes raising awareness, providing education, and advocating for legal and social changes to protect young girls and promote their well-being.

## DISCUSSION

The research paper titled "Mapping the Shadows: A Comprehensive Exploration of Breast Ironing’s Effects on Women and Girls through a Systematic Review and Scoping Analysis" provides a detailed examination of the harmful practice of breast ironing (BI). The study highlights the physical, psychological, and social consequences of BI on women and girls, while also exploring the cultural motivations and methods behind this practice

This study employed a systematic and scoping review following the PRISMA guidelines 2020 ^37^. A comprehensive search of both published and grey literature was conducted across various online databases, including EBSCO, PubMed, Google Scholar, and more. The search covered studies up to the year 2023, focusing on the biopsychosocial impacts of breast ironing.

### Psychological and Emotional Trauma

Breast ironing inflicts severe psychological and emotional trauma on young girls, causing long-lasting mental health issues such as anxiety, depression, low self-esteem, and post-traumatic stress disorder (PTSD) ^24^. Victims experience fear, shame, and social exclusion, which can impair their psychological development and social interactions ^17, 40, 49^. The inability of families to recognise these traumas exacerbates the pressure and distress within the family, particularly as the child matures ^50^.

### Pain and Discomfort

The procedure of breast ironing involves pressing, pounding, or massaging the breasts with heated or hard objects, causing immense pain and discomfort without any form of anaesthesia. This results in severe health consequences such as abscesses, infections, tissue damage, breast cancer, and psychological trauma ^7, 17^. The pain endured can be excruciating and prolonged, significantly impacting the victims’ physical and mental health.

### Inhibition of Breastfeeding

Breast ironing can severely impair a woman’s ability to breastfeed. The damage to breast tissues can make breastfeeding difficult or impossible, compromising the well-being and nutrition of future infants ^51, 52^. The long-term consequence underlines the critical impact of the practice on both maternal and child health.

### Barriers to Healthcare Access

Victims of breast ironing face numerous barriers to accessing healthcare, including stigma, fear, and lack of awareness. Many do not seek medical help due to embarrassment about their altered bodies and fear of social ostracism. Additionally, there is often a lack of trained medical personnel who can adequately address the unique needs of these victims. The social stigma and discrimination further deter families and victims from seeking medical assistance ^51^.

### Violation of Human Rights

Breast ironing is a severe violation of fundamental human rights, recognised by international bodies as a form of gender-based violence. It infringes on the physical integrity and sexual autonomy of young girls, similar to female genital mutilation (FGM). The lack of consent and the violent nature of the practice highlight its role as a tool of oppression, perpetuating gender inequality and discrimination ^5, 53^.

### Sociocultural Factors

The practice of breast ironing is deeply rooted in cultural beliefs and social norms, seen as a means to protect girls from early sexualisation, unwanted pregnancies, and societal judgment. Poverty and lack of education are major contributing factors, with traditional healers and elders endorsing the practice based on cultural beliefs ^51^. Factors such as the HIV/AIDS pandemic and urbanisation have influenced the practice, with urban-based mothers more likely to engage in breast ironing due to fears of increased exposure to sexual abuse. Addressing these sociocultural factors is crucial for designing effective interventions aimed at eradicating breast ironing.

### Impact on Sexual Health Autonomy

Breast ironing interferes with the natural sexual development of young girls, leading to distorted perceptions of sexuality and impacting their sexual self-esteem and autonomy. It denies girls the right to make decisions about their bodies and perpetuates gender inequalities, reinforcing the idea that they have limited control over their physical well-being ^49^.

### Disempowerment

Breast ironing contributes to the disempowerment of women and girls by reinforcing harmful gender norms and expectations. It strains family relationships, affects trust, and limits opportunities for education and personal development. Comprehensive efforts, including education, awareness campaigns, legal interventions, and community engagement, are essential to challenge and eliminate this harmful practice ^17^.

### Limitations

One major limitation of the study is the lack of systematic data collection and formal studies on breast ironing, particularly outside Cameroon. The reliance on self-reported data and interviews may introduce biases and limit the generalizability of the findings. Additionally, cultural sensitivities and stigma surrounding BI may result in underreporting and incomplete data.

### Implications for Practice

The findings of this study underscore the need for healthcare professionals to be aware of the signs and long-term impacts of breast ironing. This awareness is crucial for providing appropriate medical care and psychological support to survivors. Educational initiatives targeting both healthcare providers and communities are essential to raise awareness about the harmful effects of BI and to advocate for the rights of girls and women.

### Further Research

Future research should focus on collecting systematic data across different regions and populations to better understand the prevalence and impact of breast ironing. Studies should also explore the effectiveness of various interventions aimed at eradicating the practice and supporting affected individuals. Additionally, research on the role of cultural beliefs and societal norms in perpetuating BI can provide insights into designing culturally sensitive prevention programs.

### Conclusion

Breast ironing is a deeply rooted cultural practice with significant negative impacts on the physical, psychological, and social well-being of women and girls. The study highlights the urgent need for comprehensive interventions, including legal measures, community education, and support services, to protect the rights and health of affected individuals. Addressing the cultural underpinnings and societal acceptance of BI is essential for its eradication. Empowering women and girls, improving access to healthcare, and challenging sociocultural norms are critical steps towards eradicating breast ironing and ensuring the rights and health of young girls ^7, 51^. This provides a clear overview of the research paper, highlighting the key findings, health impacts, and recommendations for addressing the practice of breast ironing.

### Recommendations

1. Legal Measures: Strengthen legal frameworks to explicitly recognise and criminalise breast ironing as a form of gender-based violence.
2. Community Engagement: Implement awareness campaigns to educate communities about the harmful effects of BI and promote gender equality.
3. Support Services: Establish comprehensive support systems for survivors, including medical care, psychological counselling, and legal assistance.
4. Healthcare Training: Train healthcare providers to recognize and treat the physical and psychological consequences of BI.
5. Research and Data Collection: Conduct systematic research to gather data on the prevalence and impact of BI in various regions and evaluate the effectiveness of interventions.

## Data Availability

Relevant data underlying the research has been attached

## SUPPORTING INFORMATION

**S1 Table.**
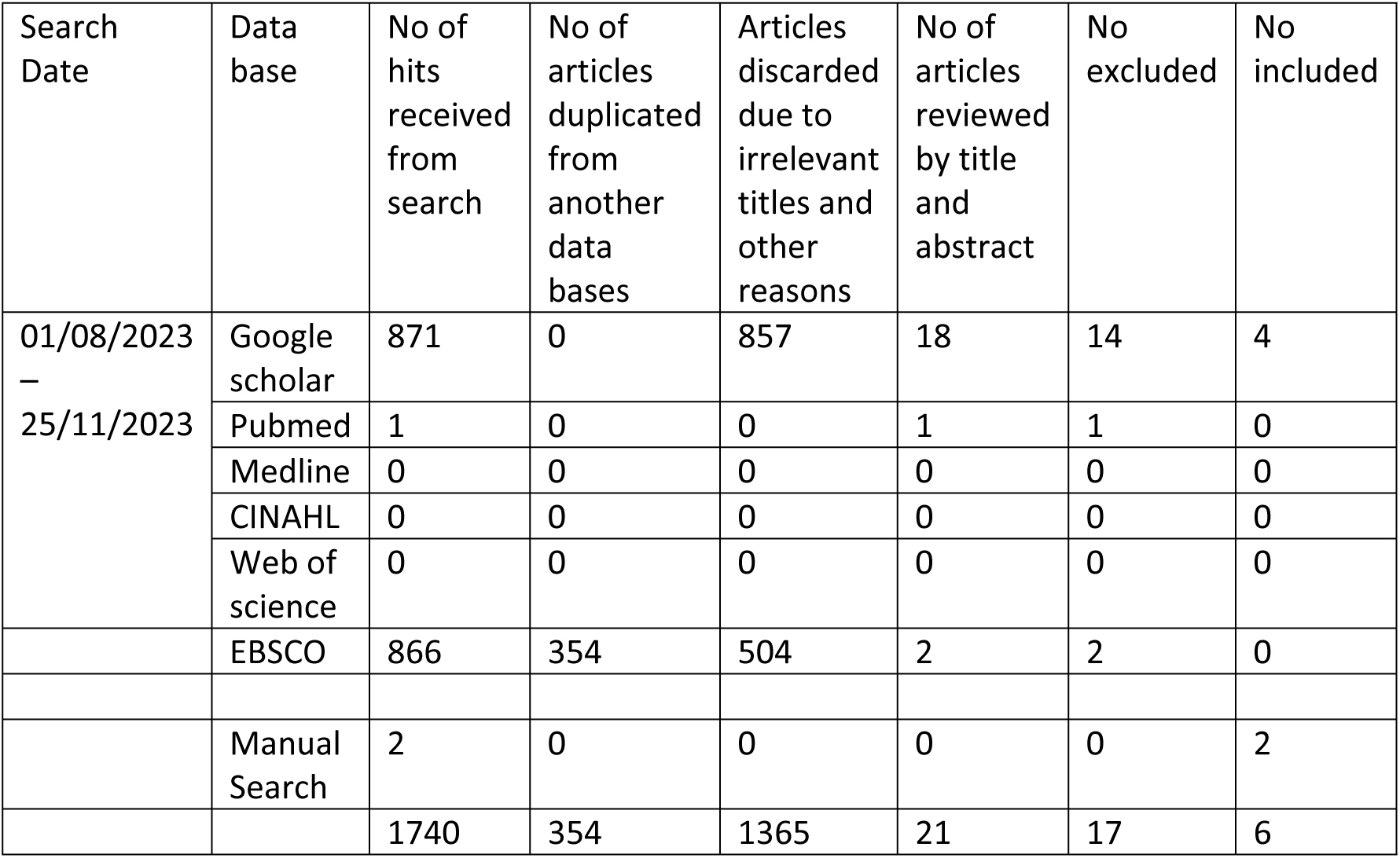

| Search Date | Data base | No of hits received from search | No of articles duplicated from another data bases | Articles discarded due to irrelevant titles and other reasons | No of articles reviewed by title and abstract | No excluded | No included |
| --- | --- | --- | --- | --- | --- | --- | --- |
| 01/08/2023 – 25/11/2023 | Google scholar | 871 | 0 | 857 | 18 | 14 | 4 |
|  | Pubmed | 1 | 0 | 0 | 1 | 1 | 0 |
|  | Medline | 0 | 0 | 0 | 0 | 0 | 0 |
|  | CINAHL | 0 | 0 | 0 | 0 | 0 | 0 |
|  | Web of science | 0 | 0 | 0 | 0 | 0 | 0 |
|  | EBSCO | 866 | 354 | 504 | 2 | 2 | 0 |
|  | Manual Search | 2 | 0 | 0 | 0 | 0 | 2 |
|  |  | 1740 | 354 | 1365 | 21 | 17 | 6 |

**S2 Table.** Boolean search.

| Date searched | Database searched | Boolean searched |
| --- | --- | --- |
| 01/08/2023<br>–<br>25/11/2023 | GOOGLE SCHOLAR | "breast ironing" OR breast flattening OR breast pounding<br>"breast ironing" OR "breast flattening" OR "breast pounding" AND "Experience"; "breast ironing" AND "breast flattening" AND "quality life" "health-related consequences and outcomes," AND "questionnaire" "interview" AND "survey"; "breast trauma" AND "breast injury" AND "harmful traditional practices against women in Africa" AND "knowledge" "attitude" AND "perceptions" AND "experiences" AND "views" AND "effects" AND "impact" AND "quality of life indicators" |
| 01/08/2023<br>–<br>25/11/2023 | PUBMED/MEDLINE | "breast ironing" AND "breast flattening" AND "breast pounding" AND "Experience"AND "Africa"; "breast ironing" OR "breast flattening" AND "Africa"; "breast ironing" AND "breast flattening" AND "breast ironing" AND "Africa" AND "experience" "health-related consequences and outcomes" AND "questionnaire" "interview," AND "survey" "breast trauma" AND "breast injury" AND "harmful traditional practices against women in Africa" AND "knowledge" "attitude" AND "perceptions" AND "experiences" AND "views" AND "effects" AND "impact" AND "quality of life indicators" |
| 01/08/2023<br>–<br>25/11/2023 | EBSCO/WEB OF SCIENCE/PSYCINFO/EMBASE | "breast pounding" AND "Experience"AND "Africa"; "breast ironing" AND "breast flattening" AND "breast pounding" AND "Experience"; "health-related consequences and outcomes" AND "questionnaire" "interview," AND "survey"; "breast trauma" AND "breast injury" AND "harmful traditional practices against women in Africa" AND "knowledge" "attitude" AND "perceptions" AND "experiences" AND "views" AND "effects" AND "impact" AND "quality of life indicators" |

**S3 Table.** Data extraction table.

| NO | Author/year | Research topic | Aims of the research | Study methodology | Sample size | COUNTRY | Findings | CRITIQUE |
| --- | --- | --- | --- | --- | --- | --- | --- | --- |
| 1 | Nkwelle N. N., (2019) | A Long-Term Health-Related Outcomes of Breast Ironing in Cameroon. | To Provide Information on the Long-Term Health Related Outcomes of Breast Ironing and its Influence on the Quality of Life of Survivors. | Quasi-Experimental Design | 230 | Cameroon | The findings of this study present positive and progressive social change implications for the individuals, families, communities, organizations and governments both local and national levels that may result in a significant alteration in behavior patterns and cultural beliefs or norms about Breast Ironing over time in Cameroon. | -The Purpose of the study was clearly stated.<br>- Relevant reviewed literature<br>-Study design appropriate<br>-Informed consent stated<br>- well presented data and analysis of results<br>**Overall-** good |
| 2 | Nyangweso M., (2022) | Contending with Health Outcomes of Sanctioned Rituals: The Case of Puberty Rites. | This paper explores the rites of passage rituals as the loci of health outcomes. It highlights how religiously sanctioned practices play a central role in healthcare in defiance of the perceived private and public dichotomy that dominates the modern secular mindset. Highlighted in the chapter are African rites of passage, specifically breast<br>“ironing”, female<br>genital<br>mutilation/cutting<br>(FGM/C), and<br>child marriage. | The study employed a cross-sectional intersectionality methodology that drew upon a mixed research investigative approach, namely a qualitative and quantitative research design. | 60 | Kenya | Drawing from findings of a survey of 50 respondents, the chapter illustrates how these practices exemplify how rituals invoke health concerns in Africa and amongst Africans in the diaspora. | -The Purpose and Aim of the study were not clearly stated.<br>- Relevant reviewed literature<br>-Study design appropriate<br>-Informed consent stated<br>-The data were not clearly stated, not well presented, and lack some organizational structure. Analysis of results not clearly stated.<br>**Overall-** poor. |

| NO | Author/year | Research topic | Aims the research | Study methodology | Sample size | COUNTRY | Findings | CRITIQUE |
| --- | --- | --- | --- | --- | --- | --- | --- | --- |
| 3. | Pemunta, N.V., 2016 | The Social Context of Breast Ironing in Cameroon. | To explore the confluence of social factors and the social context underpinning the pervasive practice of breast ironing in Cameroon. | Qualitative Research | 27<br>(16 girls submitted to BI, 5 activists, 6 mothers who had administered breast ironing on their daughters.) | Cameroon | The social phenomenon of breast ironing is a disciplinary technique meant to conserve the social body politic and to ensure the wellbeing of the girl child. | - Purpose stated obviously<br>- Appropriate literature reviewed<br>- Informed consent was stated<br>- Well presented data and analysis.<br>- Overall - Good |
| 4 | Tapscott, R., 2012 | Understanding Breast “Ironing”: A Study of the methods, motivations, and outcomes of breast flattening practices in Cameroon | To better understand the practice of breast flattening, including how, when, where, and why it is practiced. | Qualitative Research | 75 | Cameroon | Breast flattening is a painful practice considered the norm for many girls who experience it. | - Purpose stated partially<br>- Appropriate literature reviewed<br>- Informed consent was not stated<br>- Poor presented data and analysis.<br>- Overall - Fair |
| 5 | Falana, 2020 | Breast Ironing: a rape of the girl-child’s Personality integrity and sexual autonomy | The focus of this paper is to determine whether the practice of breast ironing could be rightly considered as a cultural | Qualitative Research approach and the Expository analytic method | None | Unknown | The findings of this research therefore, it is germane to argue that the harrowing practice of breast ironing is a rape of the victim’s personality integrity, personal | - Purpose stated obviously<br>- lack of appropriate literature reviewed<br>- Informed consent was not stated |
|  |  |  | practice that is imperative (essential); or whether it could be ideally seen as an unnecessary and callous cultural practice that constitutes a brute and violent violation of the girl child's right to sexual autonomy and right to physical integrity. |  |  |  | liberty, and the right to sexual autonomy. | - Poor presented data and analysis.<br>- Overall - Poor |
| 6 | Oniyangi and Tosin, 2022 | Influence of Cultural Practices on the Health of Yoruba People Living in Ilorin South LGA, Kwara State | Examines the health impacts of cultural practices such as breast ironing, puerperal baths, and | Descriptive research design using survey method | 200 | Kwara state, Nigeria | Breast Ironing<br>- Negative Influence: 88.6% of respondents acknowledged negative health impacts.<br>- Health Effects: Causes swelling, | - Purpose stated<br>- Appropriate literature reviewed<br>- Ethical clearance taking from the university but |
|  |  |  | nutritional taboos among the Yoruba people |  |  |  | burning, irritation, increased risk of breast cancer, fever, extreme pain, and delay in breast growth.<br>- Chi-Square Analysis: Calculated value of 532.24 (greater than critical value of 16.92), confirming significant negative influence.<br>Other results focussed on puerperal bath and Nutritional Taboos | Informed consent was stated stated<br>- Well presented data and analysis including frequency counts, percentages and chi-square tests at 0.05 alpha level..<br>- Overall - Good |

**S4 Table.** ELIGIBILITY OF SELECTED RCT ARTICLES USING TULDER’S (2003) ASSESSMENT TOOL.

| Authors | Was the method of randomization adequate? | Was the treatment allocation concealed? | Were the groups similar at baseline regarding the most prognostic factor? | Was the patient blinded to the intervention? | Was the care provider blinded to the intervention? | Was the outcome assessor blinded to the intervention? | Were co-interventions avoided or similar? Was the compliance acceptable in all groups? | Was the compliance acceptable in all groups? | Was the dropout rate described and acceptable? | Was the timing of the outcome assessment in both groups comparable? | Did the study include an intention to treat analysis? | TOTAL |
| --- | --- | --- | --- | --- | --- | --- | --- | --- | --- | --- | --- | --- |
| 1.Nkwelle<br>N. N. N.,<br>(2019) | Yes | NO | Yes | NO | NO | NO | NO | NO | YES | Yes | NO | 4/11 |

**S5 Table.**
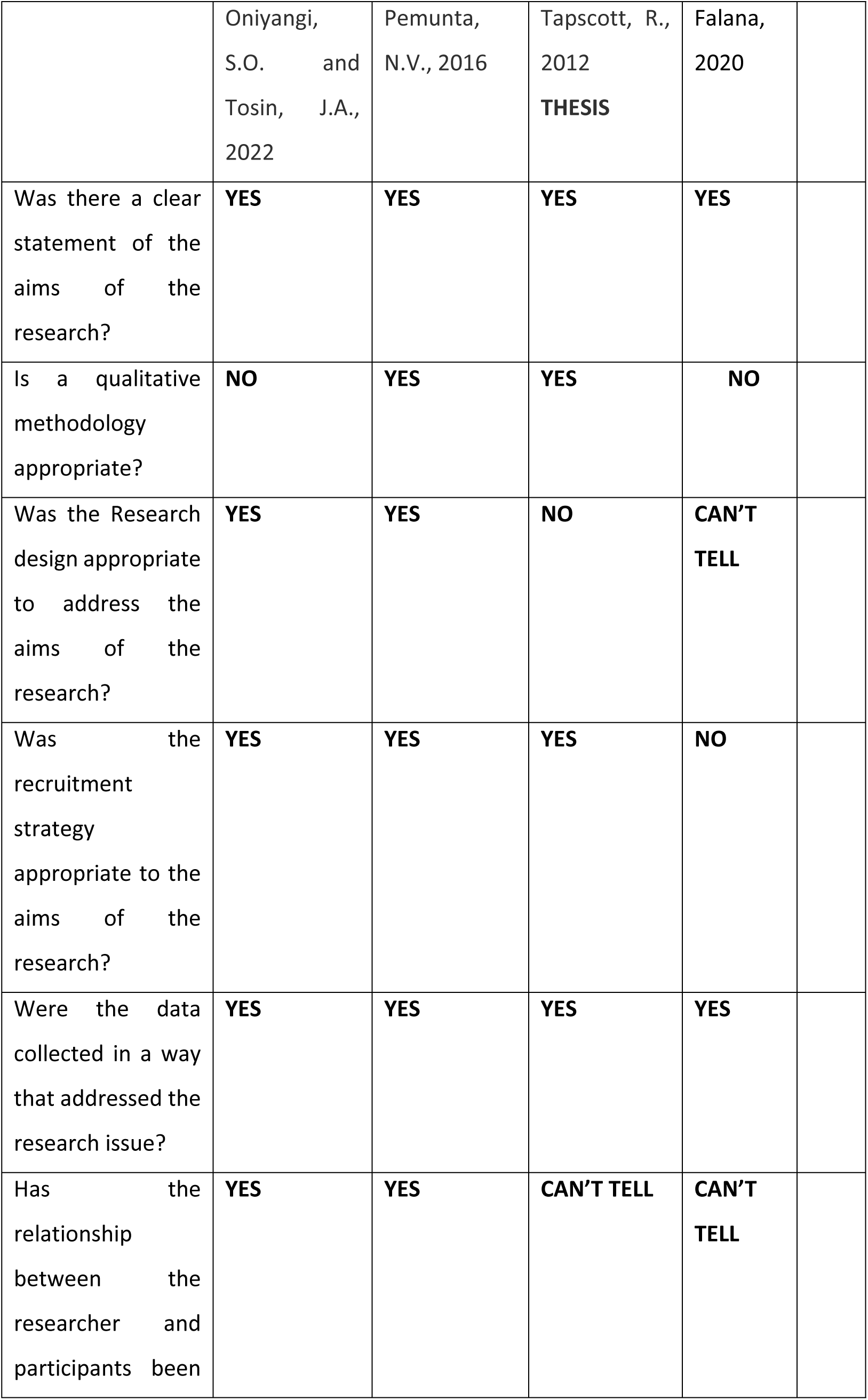
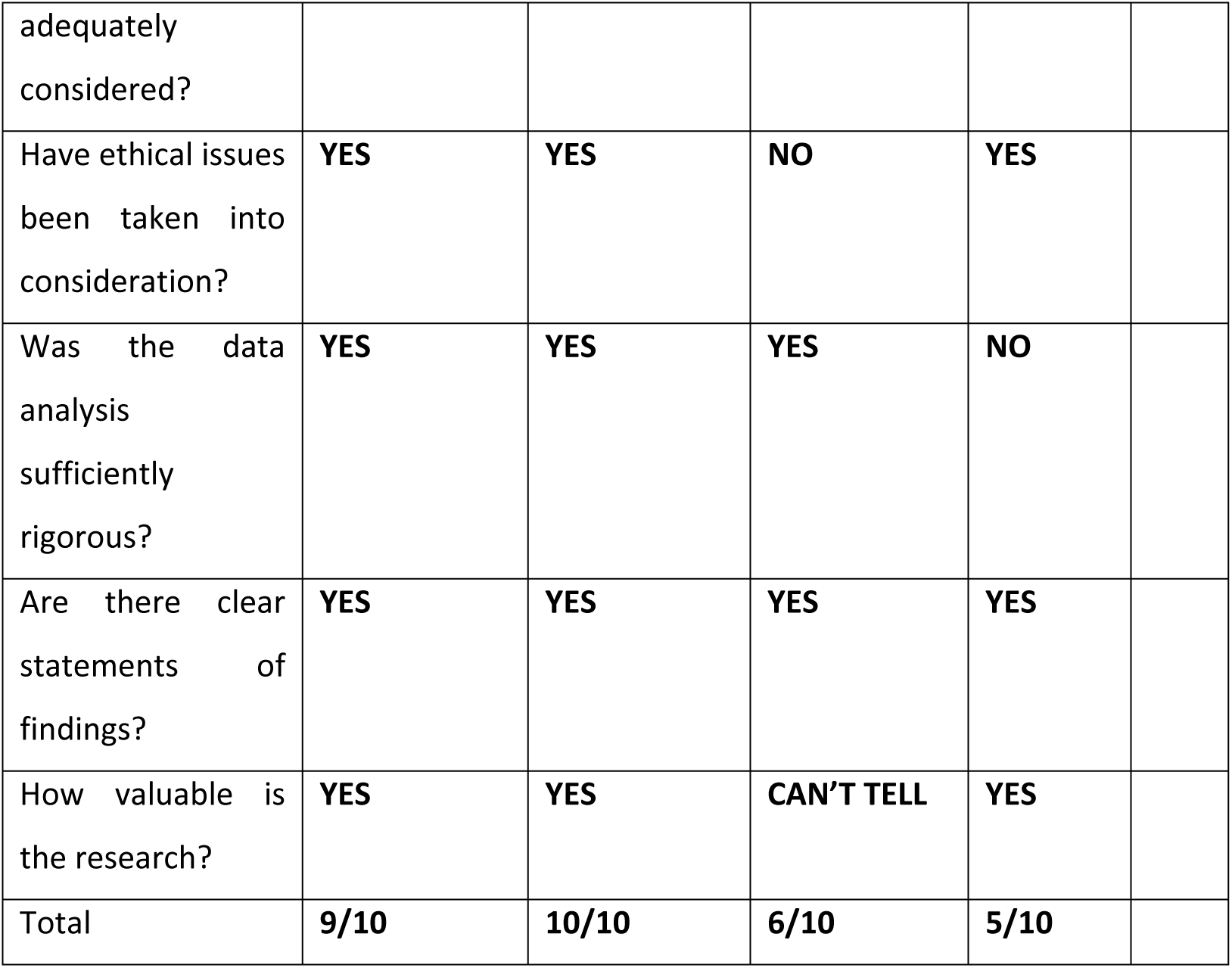
- CASP QUESTIONS FOR QUALITATIVE STUDIES (Public Health Resource Unit E*n*gland, 2006)

**S6 Table.** Mixed Methods Appraisal Tool (MMAT), version 2018 -.

| Category of study designs | Methodological quality criteria | Nyangweso M., (2022) |  |  |
| --- | --- | --- | --- | --- |
|  |  | Yes | No | Can't tell |
| Screening questions (for all types) | S1. Are there clear research questions? |  | NO |  |
|  | S2. Do the collected data allow to address the research questions? |  | NO |  |
| 1. Qualitative | 1.1. Is the qualitative approach appropriate to answer the research question? |  | NO |  |
|  | 1.2. Are the qualitative data collection methods adequate to address the research question? |  | NO |  |
|  | 1.3. Are the findings adequately derived from the data? |  | NO |  |
|  | 1.4. Is the interpretation of results sufficiently substantiated by data? |  | NO |  |
|  | 1.5. Is there coherence between qualitative data sources, collection, analysis and interpretation? |  | NO |  |
| 2. Quantitative randomized controlled trials | 2.1. Is randomization appropriately performed? |  | NO |  |
|  | 2.2. Are the groups comparable at baseline? |  | NO |  |
|  | 2.3. Are there complete outcome data? |  | NO |  |
|  | 2.4. Are outcome assessors blinded to the intervention provided? |  | NO |  |
|  | 2.5. Did the participants adhere to the assigned intervention? |  | NO |  |
| 3. Quantitative non-randomized | 3.1. Are the participants representative of the target population? |  | NO |  |
|  | 3.2. Are measurements appropriate regarding both the outcome and intervention (or exposure)? |  | NO |  |
|  | 3.3. Are there complete outcome data? |  | NO |  |
|  | 3.4. Are the confounders accounted for in the design and analysis? | YES | NO |  |
|  | 3.5. During the study period, is the intervention administered (or exposure occurred) as intended? |  | NO |  |
| 4. Quantitative descriptive | 4.1. Is the sampling strategy relevant to address the research question? |  | NO |  |
|  | 4.2. Is the sample representative of the target population? |  | NO |  |
|  | 4.3. Are the measurements appropriate? |  | NO |  |
|  | 4.4. Is the risk of nonresponse bias low? |  | NO |  |
|  | 4.5. Is the statistical analysis appropriate to answer the research question? |  | NO |  |
| 5. Mixed methods | 5.1. Is there an adequate rationale for using a mixed methods design to address the research question? | YES |  |  |
|  | 5.2. Are the different components of the study effectively integrated to answer the research question? | YES |  |  |
|  | 5.3. Are the outputs of the integration of qualitative and quantitative components adequately interpreted? |  | NO |  |
|  | 5.4. Are divergences and inconsistencies between quantitative and qualitative results adequately addressed? |  | NO |  |
|  | 5.5. Do the different components of the study adhere to the quality criteria of each tradition of the methods involved? | YES |  |  |
| Total |  | Y-4 | 24 | 0 |

\*Consider, if feasible to do so, reporting the number of records identified from each database or register searched (rather than the total number across all databases/registers).
\*\*All records were excluded by a human and none was excluded by automation tools.

